# A randomized open-label, observational study of the novel ketone ester, bis octanoyl (R)-1,3-butanediol, and its acute effect on ß-hydroxybutyrate and glucose concentrations in healthy older adults

**DOI:** 10.1101/2024.04.16.24305925

**Authors:** Elizabeth B. Stephens, Chatura Senadheera, Stephanie Roa-Diaz, Sawyer Peralta, Laura Alexander, Wendie Silverman-Martin, Michi Yukawa, Jenifer Morris, James B. Johnson, John C. Newman, Brianna J. Stubbs

**Affiliations:** Buck Institute for Research on Aging, Novato, CA, USA; Veteran’s Affairs Medical Center, San Francisco, CA, USA; Independent Researcher, Greenbrae, CA, USA; Division of Geriatrics, University of California, San Francisco, CA, USA

**Keywords:** ketones, ketone ester, ketone di-ester, exogenous ketone, beta-hydroxybutyrate

## Abstract

Bis-octanoyl (R)-1,3-butanediol (BO-BD) is a novel ketone ester (KE) ingredient which increases blood beta-hydroxybutyrate (BHB) concentrations rapidly after ingestion. KE is hypothesized to have beneficial metabolic effects on health and performance, especially in older adults. Whilst many studies have investigated the ketogenic effect of KE in young adults, they have not been studied in an exclusively older adult population, for whom age-related differences in body composition and metabolism may alter the effects. This randomized, observational, open-label study in healthy older adults (n = 30, 50% male, age = 76.5 years, BMI = 25.2 kg/m^2^) aimed to elucidate acute tolerance, blood BHB and blood glucose concentrations for 4 hours following consumption of either 12.5 or 25 g of BO-BD formulated firstly as a ready-to-drink beverage (n = 30), then as a re-constituted powder (n = 21), taken with a standard meal. Both serving sizes and formulations of BO-BD were well tolerated, and increased blood BHB, inducing nutritional ketosis (≥ 0.5mM) that lasted until the end of the study. Ketosis was dose responsive; peak BHB concentration (C_max_) and incremental area under the curve (iAUC) were significantly greater with 25 g compared to 12.5 g of BO-BD in both formulations. There were no significant differences in C_max_ or iAUC between formulations. Blood glucose increased in all conditions following the meal; there were no consistent significant differences in glucose response between conditions. These results demonstrate that both powder and beverage formulations of the novel KE, BO-BD, induce ketosis in healthy older adults, facilitating future research on functional effects of this ingredient in aging.

## Introduction

Beta-hydroxybutyrate (BHB), acetoacetate, and acetone are naturally occurring molecules derived from fats, known collectively as ketone bodies (or ketones). Ketones serve two key functions in mammalian biology. First, ketones act as a metabolic fuel source, enabling the conversion of stored energy from lipids into a readily usable form by the brain, heart, and peripheral tissues, extending survival during periods of starvation, such as fasting [1]. Second, ketones play a role as signaling molecules within an environmentally responsive network that regulates both health span and lifespan in model organisms [2]. The signaling effects of BHB have been detected at blood concentrations as low as 0.2 mM [3]. However, a state of ketosis is more commonly defined as blood BHB concentrations at, or exceeding 0.5 mM, 5-10-fold higher than constitutive blood BHB levels [4–6]. Physiological ketosis typically arises when dietary carbohydrate intake is restricted, such as during fasting or following a very low-carbohydrate, low fat diet [7, 8]. In contrast, pathological ketoacidosis, characterized by blood BHB levels exceeding 10 mM, is seen during states of metabolic dysregulation, such as alcoholic ketoacidosis and in poorly controlled type 1 diabetes [9].

Endogenous ketosis involves lipolysis of adipose tissue, providing the liver with fatty acids for ketone production [10]. By contrast, exogenous ketosis can occur following the consumption of dietary sources of ketones, such as ketone esters (KE). Exogenous ketones allow the elevation of blood ketone levels without substantial changes in diet or macronutrient intake. Although exogenous ketone consumption can mimic certain aspects of endogenous ketosis, the extent to which it replicates all the biological effects of natural ketosis in humans remains uncertain [6, 11]. KE have been employed to investigate the impact of exogenous ketosis on various aspects of health and disease, ranging from physical and cognitive performance to blood glucose control and cardiac function [12–24].

Ketosis is a feature of metabolic states that are associated with improved healthspan and lifespan in preclinical models, including caloric restriction [25, 26], ketogenic diets [27, 28] and direct treatment with BHB [29]. Signaling functions of BHB modulate multiple ‘hallmarks of aging’ *in vitro* and *in vivo*, including inflammation [30, 31], senescence [32], mitochondrial function, epigenetics [33, 34] and proteostasis [35]. Therefore, there is growing interest in exogenous ketones as a strategy to conveniently deliver ketosis in older adult populations to target these mechanisms of aging. However, whilst extensive studies of younger adults exist, no studies to date have examined if KE are tolerable and efficacious in an exclusively older adult population. Preclinical studies suggest that metabolism of endogenous and exogenous ketones could change with age [36, 37]. Furthermore, given the differences in body composition, metabolism of key organs such as the liver and kidney, increased use of concomitant medications and reduced functional reserve that may be present in an older adult population [38, 39], it is critical to explore the basic metabolic effects and tolerance of KE in healthy older adults before progressing to studies in populations with functional deficits.

Bis-octanoyl (R)-1,3-butanediol (BO-BD) is a novel fatty-acid ketone ester that robustly stimulates hepatic ketone production, independently of dietary carbohydrate intake [40]. Rapid enzymatic breakdown of fatty-acid ketone esters within the small intestine results in the formation of ketogenic precursors: medium-chain fatty acids and (R)-1,3-butanediol–an alcohol with ketogenic properties [41]. These breakdown products are transported to the liver through the portal circulation. Medium-chain fatty acids serve as a foundational substrate for ‘classical’ hepatic ketogenesis, leading to the release of BHB and acetoacetate [42]. Meanwhile, (R)-1,3-butanediol undergoes a ‘non-classical’ hepatic ketogenic pathway, converting into BHB [43, 44]. When orally administered to rats, mice, and adult humans, BO-BD and the closely related fatty-acid KE, bis-hexanoyl (R)-1,3-butanediol (BH-BD), deliver rapid increases in blood BHB concentrations that remain elevated for several hours [41, 45, 46].

Data on the ketogenic effect of BO-BD in humans is limited, with only one publication to date that demonstrated equivalent metabolism between matched servings of BO-BD and BH-BD [40]. Furthermore, a novel powdered formulation of BO-BD recently became available; whilst previous work demonstrated equivalent ketogenic potential of both beverage and powdered formulations of BH-BD [40], there is no kinetic data on the powdered form of BO-BD. To address the limited kinetic data for different formulations and serving sizes of BO-BD and the lack of data on KE metabolism in older adults, we undertook a randomized, parallel group, open-label, observational study in healthy adults, aged 65 or older, to determine the blood BHB and glucose kinetics of two serving sizes of BO-BD beverage (n = 30), as well as tolerability. This kinetics investigation was carried out as the first visit of a longer study designed to investigate the safety and tolerability of BO-BD taken for 12-weeks in older adults (NCT05585762), which is described in a protocol paper pre-print [47]. The powdered BO-BD formulation became available during the 12-week study, so subjects were invited to re-consent and repeat the kinetics testing with BO-BD powder (n = 21). We hypothesized that BO-BD would be well tolerated and would induce nutritional ketosis (BHB ≥ 0.5 mM) in a serving-size dependent manner in healthy older adults.

## Materials and Methods

### Study overview

The data presented here was collected during the ‘acute ketone kinetics visits’ of a 12-week study designed to address the safety and tolerability of BO-BD in healthy older adults (NCT05585762), carried out in the Clinical Research Unit at The Buck Institute for Research on Aging (Novato, CA, USA) [47]. Healthy adults at or over the age of 65 (n = 30) took part in a randomized, parallel-group, observational, open-label study, in which capillary blood BHB and glucose concentrations were measured before and for 4 hours following ingestion of 12.5 and 25 g serving sizes of BO-BD (common name, C8 ketone diester) formulated in a beverage. An external study statistician prepared a 4-block randomization sequence stratified by sex, which specified serving size (12.5 or 25 g) as well as intervention for the main 12-week study (BO-BD or placebo; this phase of the study will be reported separately), and participants were allocated a randomization number by study staff based on order of enrollment. Study staff and participants were not blinded to their serving size allocation for the acute ketone kinetics visit. During the study, a new powdered form of BO-BD that can be reconstituted in water became available. Therefore, we amended our protocol to include an optional second acute ketone kinetics visit, where subjects who completed the 12-week study and who chose to re-consent (n = 21) returned and completed a cross-over test, where they consumed the same BO-BD serving size allocation as their first visit, but with the powder study product (**Figure 1**). The study was approved by Advarra IRB on September 28^th^ 2022; and randomized the first subject on January 31^st^ 2023. The amendment to add the powder formulation was approved on October 18^th^ 2023, and the final subject completed testing on the powder formulation on January 17^th^ 2024, which formally ended the study. The study was conducted in accordance with the Declaration of Helsinki ^35^.

**Figure 1.**
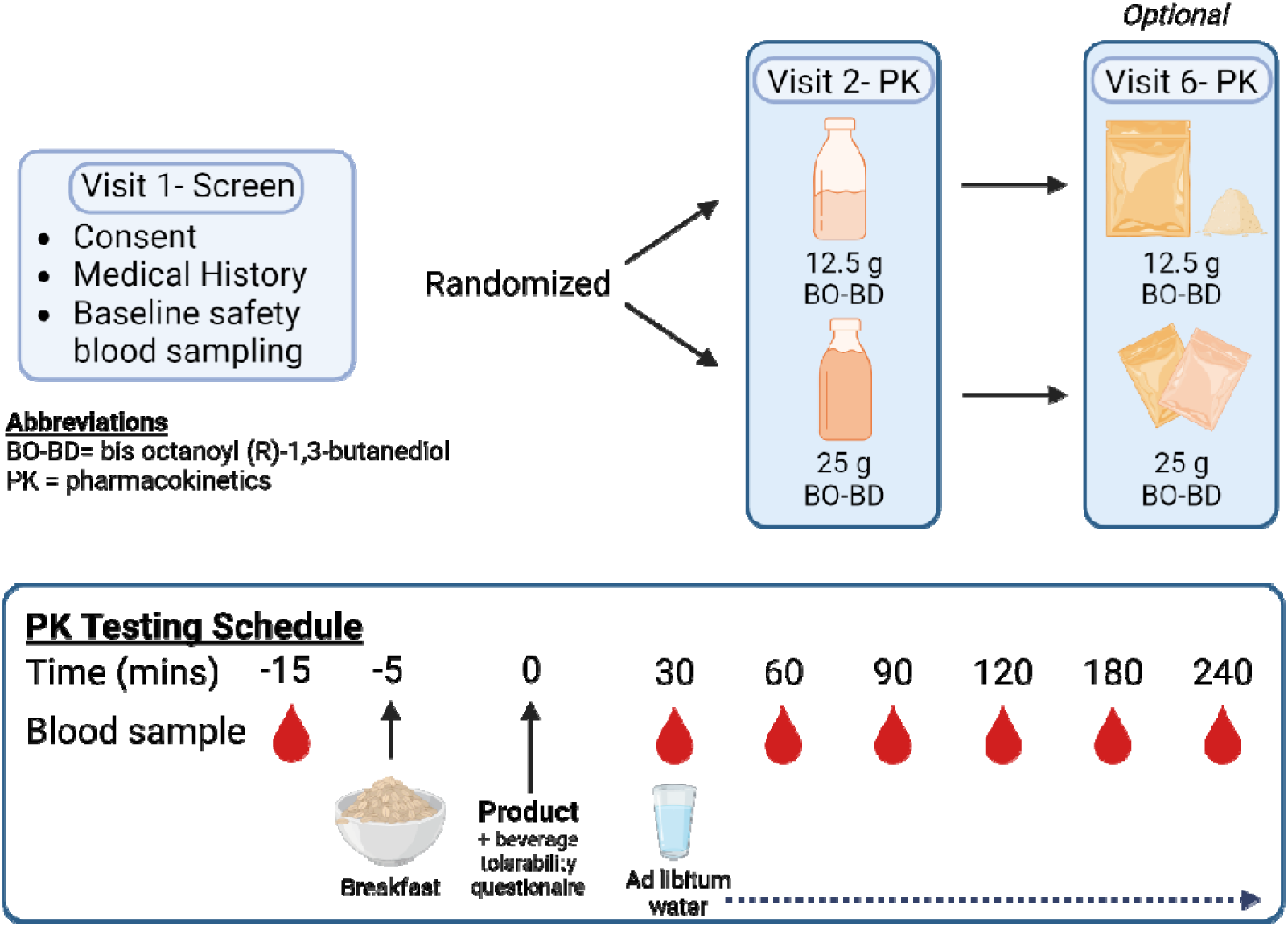
Schematic showing overall study design, study products and servings, and schedule for pharmacokinetics (PK) visits. Figure created using BioRender.com.

### Participants and screening

After providing an explanation of the study and requirements, participants provided informed consent if they remained interested in participating and prior to inclusion. Interested participants completed a medical history interview to determine eligibility. Participants included healthy older adults with stable chronic disease who live independently in the community and have no significant functional impairments (≥ 65 years old, BMI 18.5 – 34.9 kg/m^2^, male, n = 15, beverage 12.5 g = 8, 25 g = 7; female, n = 15, beverage 12.5 g = 7, 25 g = 8), a sub-set of these (total n = 21; male n = 12, powder 12.5 g = 7, 25 g = 5; female, n = 9, powder 12.5 g = 5, 25 g = 4) re-consented to consume the powder product. Subject characteristics are shown in **Table 1**. To complete the optional powder acute ketone kinetics visit, subjects were required to have completed the 12-week study, and to provide updated consent for the optional visit. Exclusion criteria included current clinically important or unstable illness, clinically important gastrointestinal conditions, allergies to ingredients in the test products, unstable use of medications or supplements, non-ambulation, a Canadian Study of Health and Aging clinical frailty score greater than 5, requiring assistance with any activity of daily living (excluding continence), cognitive impairment sufficient to prevent informed consent, living in an institutional setting, insufficient fluency in English to provide informed consent, following a ketogenic diet or using ketone supplements, and pregnancy, lactation, or the possibility of becoming pregnant during the study.

**Table 1.**
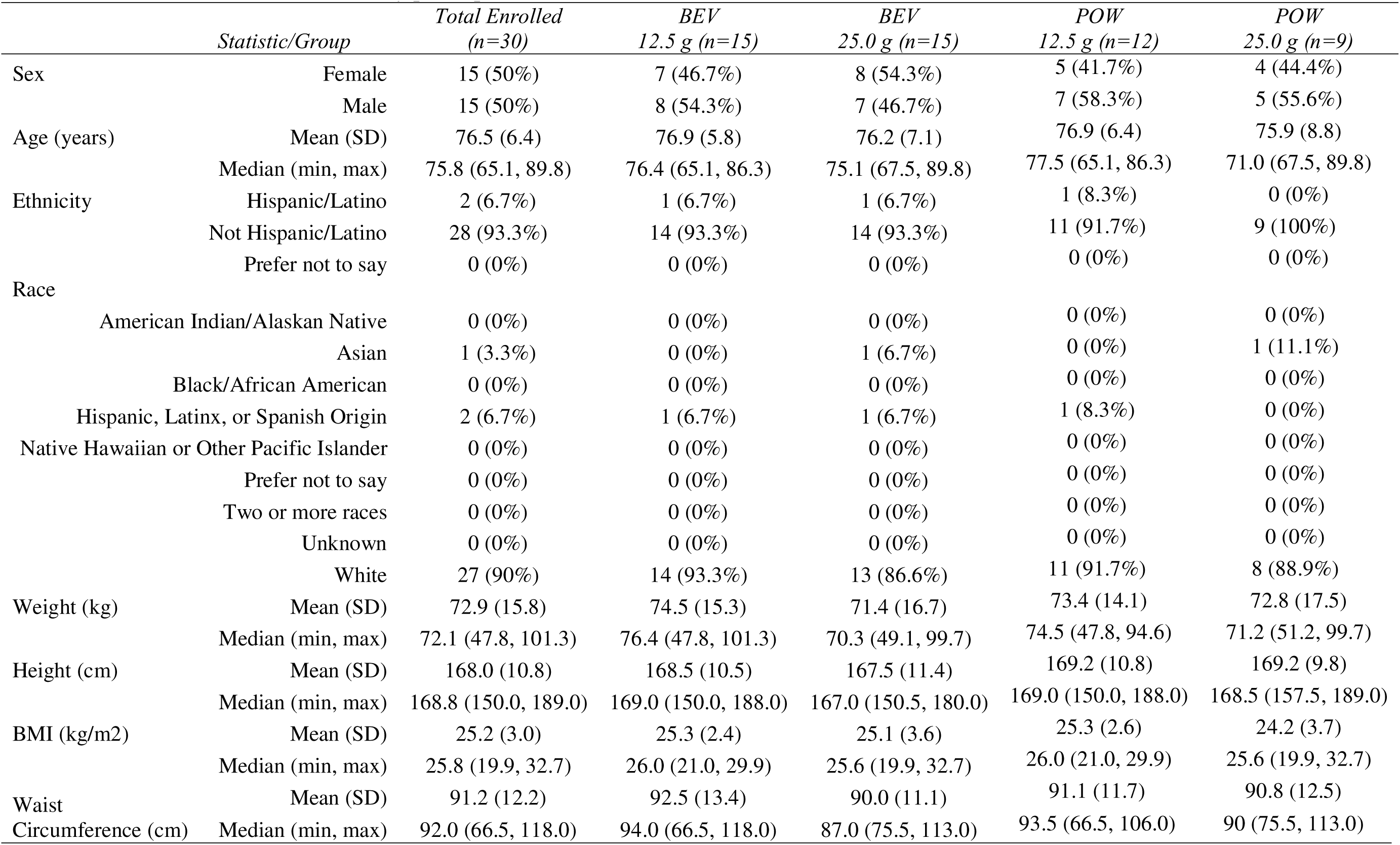
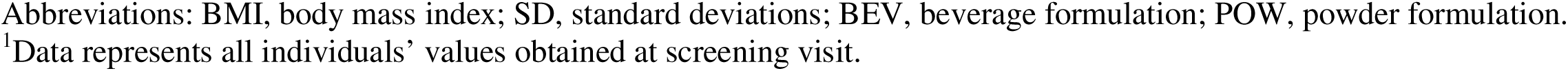
Baseline characteristics of study participants.

| *Statistic/Group* |  | *Total Enrolled*<br>*(n=30)* | *BEV*<br>*12.5 g (n=15)* | *BEV*<br>*25.0 g (n=15)* | *POW*<br>*12.5 g (n=12)* | *POW*<br>*25.0 g (n=9)* |
| --- | --- | --- | --- | --- | --- | --- |
| Sex | Female | 15 (50%) | 7 (46.7%) | 8 (54.3%) | 5 (41.7%) | 4 (44.4%) |
|  | Male | 15 (50%) | 8 (54.3%) | 7 (46.7%) | 7 (58.3%) | 5 (55.6%) |
| Age (years) | Mean (SD) | 76.5 (6.4) | 76.9 (5.8) | 76.2 (7.1) | 76.9 (6.4) | 75.9 (8.8) |
|  | Median (min, max) | 75.8 (65.1, 89.8) | 76.4 (65.1, 86.3) | 75.1 (67.5, 89.8) | 77.5 (65.1, 86.3) | 71.0 (67.5, 89.8) |
| Ethnicity | Hispanic/Latino | 2 (6.7%) | 1 (6.7%) | 1 (6.7%) | 1 (8.3%) | 0 (0%) |
|  | Not Hispanic/Latino | 28 (93.3%) | 14 (93.3%) | 14 (93.3%) | 11 (91.7%) | 9 (100%) |
|  | Prefer not to say | 0 (0%) | 0 (0%) | 0 (0%) | 0 (0%) | 0 (0%) |
| Race | American Indian/Alaskan Native | 0 (0%) | 0 (0%) | 0 (0%) | 0 (0%) | 0 (0%) |
|  | Asian | 1 (3.3%) | 0 (0%) | 1 (6.7%) | 0 (0%) | 1 (11.1%) |
|  | Black/African American | 0 (0%) | 0 (0%) | 0 (0%) | 0 (0%) | 0 (0%) |
|  | Hispanic, Latinx, or Spanish Origin | 2 (6.7%) | 1 (6.7%) | 1 (6.7%) | 1 (8.3%) | 0 (0%) |
|  | Native Hawaiian or Other Pacific Islander | 0 (0%) | 0 (0%) | 0 (0%) | 0 (0%) | 0 (0%) |
|  | Prefer not to say | 0 (0%) | 0 (0%) | 0 (0%) | 0 (0%) | 0 (0%) |
|  | Two or more races | 0 (0%) | 0 (0%) | 0 (0%) | 0 (0%) | 0 (0%) |
|  | Unknown | 0 (0%) | 0 (0%) | 0 (0%) | 0 (0%) | 0 (0%) |
|  | White | 27 (90%) | 14 (93.3%) | 13 (86.6%) | 11 (91.7%) | 8 (88.9%) |
| Weight (kg) | Mean (SD) | 72.9 (15.8) | 74.5 (15.3) | 71.4 (16.7) | 73.4 (14.1) | 72.8 (17.5) |
|  | Median (min, max) | 72.1 (47.8, 101.3) | 76.4 (47.8, 101.3) | 70.3 (49.1, 99.7) | 74.5 (47.8, 94.6) | 71.2 (51.2, 99.7) |
| Height (cm) | Mean (SD) | 168.0 (10.8) | 168.5 (10.5) | 167.5 (11.4) | 169.2 (10.8) | 169.2 (9.8) |
|  | Median (min, max) | 168.8 (150.0, 189.0) | 169.0 (150.0, 188.0) | 167.0 (150.5, 180.0) | 169.0 (150.0, 188.0) | 168.5 (157.5, 189.0) |
| BMI (kg/m2) | Mean (SD) | 25.2 (3.0) | 25.3 (2.4) | 25.1 (3.6) | 25.3 (2.6) | 24.2 (3.7) |
|  | Median (min, max) | 25.8 (19.9, 32.7) | 26.0 (21.0, 29.9) | 25.6 (19.9, 32.7) | 26.0 (21.0, 29.9) | 25.6 (19.9, 32.7) |
| Waist<br>Circumference (cm) | Mean (SD) | 91.2 (12.2) | 92.5 (13.4) | 90.0 (11.1) | 91.1 (11.7) | 90.8 (12.5) |
|  | Median (min, max) | 92.0 (66.5, 118.0) | 94.0 (66.5, 118.0) | 87.0 (75.5, 113.0) | 93.5 (66.5, 106.0) | 90 (75.5, 113.0) |

Abbreviations: BMI, body mass index; SD, standard deviations; BEV, beverage formulation; POW, powder formulation.
<sup>1</sup>Data represents all individuals' values obtained at screening visit.

### Study products

BO-BD was formulated into a tropical flavored beverage (water, high fat whey protein concentrate, modified gum acacia, citric acid, soy lecithin, natural flavors, stevia leaf extract, pectin, sodium carboxymethyl cellulose, potassium sorbate) packaged into 75 mL bottles containing 25 g BO-B, or an unflavored powder (soluble corn fiber, sodium caseinate) mixed into 8 – 12 fl oz of water using a shaker. Study products were provided by BHB Therapeutics Ltd (Dublin, Ireland), conflict of interest management was undertaken as described in the methods preprint [47]. On the day of the test, investigators administered either 12.5 g BO-BD (37.5 mL beverage, or 8 fl oz water + 19 g powder) or 25 g BO-BD (75 mL beverage, or 12 fl oz water + 38 g powder). Participants were randomly allocated their serving size based on a block randomization sequence (block size = 4) that resulted in a 1:1 breakdown of subjects to receive either 12.5 g or 25 g of ketone ester, split evenly between male and female participants for the beverage testing. For the powder testing the split, maintaining the prior dose assignment, was as follows: 12.5 g male = 7, female = 5; 25 g male = 5, female = 4. Study product nutritional information is shown in **Table 2**.

**Table 2.** Nutritional information of Study Beverage and Standard Meal.

|  | BEV | BEV | POW | POW | Standard |
| --- | --- | --- | --- | --- | --- |
|  | 12.5 g | 25 g | 12.5 g | 25 g | Meal |
| BO-BD (g) | 12.5 | 25 | 12.5 | 25 | - |
| Energy (kcal) | 120 | 240 | 120 | 240 | 280 |
| Carbohydrate (g) | 1 | 2 | 3 | 6 | 24 |
| Fat (g) | 0.25 | 0.5 | 0 | 0 | 14 |
| Protein (g) | 1 | 2 | 2 | 4 | 17 |
| Fiber (g) | 0 | 0 | 3 | 3 | 5 |

**Abbreviations:** BO-BD, bis octanoyl (R)-1,3 butanediol; BEV, beverage formulation; POW, powder formulation.

### Standard meal

The standardized meal consumed before study product administration on all test days was a sachet of instant oatmeal (Nature’s Path Organic Blaine, WA, USA – Gluten Free Homestyle Instant Oatmeal), prepared with water and a half serving of whey protein powder (Momentous, Menlo Park, CA, USA – Essential Protein, Grass Fed Whey Isolate, Vanilla Flavor) Nutritional information for the meal is shown in **Table 2**.

### Study procedures

Participants completed one in-person consent and screening meeting with an investigator prior to the first test day. The meeting involved explanation of the study procedures and informed consent, preliminary blood and urine tests, and a medical history interview to determine eligibility. On study test days, participants arrived at the testing facility fasted for 10 h, having consumed no alcohol for 10 h, completed no exercise for 10 h and with no acute illness. Participants confirmed that they met these requirements by responding to interview questions by the investigator and completed a series of questions to assess baseline presence of gastrointestinal (GI) and systemic symptoms as previously described [45]. Briefly, the questionnaire features 10 symptoms (gas, nausea, vomiting, abdominal cramping, stomach rumbling, burping, reflux, diarrhea, headache, dizziness) that are scored none (0), mild (1), moderate (2) or severe (3), leading to a maximal score of 30. Investigators then obtained the participants’ fasted BHB and glucose measurements prior to consumption of the study breakfast. Immediately after breakfast consumption, participants were given and instructed to drink their allocated serving of study product. Investigators obtained BHB and glucose measurements at 30, 60, 90, 120, 180, and 240 minutes after study product consumption (**Figure 1**).

BHB and glucose measurements were measured using a finger stick with a lancet to obtain capillary blood for testing using a blood glucose and beta-ketone dual monitoring system (KetoMojo, Napa, USA). Before each finger stick, the participants finger was cleaned with an alcohol wipe and allowed to air dry before lancing. The first drop of blood was wiped away and the following drops of blood were used for blood glucose and ketone concentration measurements.

Participants were asked to remain sedentary throughout the duration of the test and not to consume food or caloric beverages; non-caloric beverages (e.g., tea or water) were permitted *ad libitum*. After the last BHB and glucose measurement, participants repeated the symptom questionnaire to assess any changes that occurred after the standard breakfast and study product, and adverse events were assessed using an open-ended question. Data input sheets were manually checked for possible errors, omissions, or protocol deviations prior to database entry.

### Statistical methods

As this investigation was carried out as part of a larger study with the primary outcome of safety and tolerance, there was no power calculation carried out to determine the sample size for this sub-study. Our primary outcome of interest was changes in BHB concentrations between serving sizes and between formulations, specifically maximal BHB concentration (BHB C_max_) at any point in the 4h test days. Based on previous studies with BO-BD, which found a BHB C_max_ of [mean (SD)] 0.98 (0.2) mM in 12.5g and 1.76 (0.5) mM with 25 g servings, our sample size of n = 15 per group provides 98% power to detect an effect of the same size with a two-sided alpha of 0.05.

All subjects who consumed study product completed the 4h test day, so reported data is from all subjects who received the study products. BHB kinetics were analyzed using the raw data. BHB C_max_ and BHB T_max_ were manually extracted from the raw data (secondary outcomes). Planned two-way comparisons were made between the two serving sizes of the same formulation (i.e., 12.5 vs 25 g) using data from all available subjects (beverage n = 30, powder n = 21), and between matched serving sizes of the two formulations (i.e., beverage vs. powder) using only data from subjects who completed the cross-over visit (n = 21), after we established that there were no significant differences in the demographics of the populations who completed one or both visits. Exploratory analysis included correlations between body weight and BHB kinetic parameters and testing for sex-differences in BHB kinetic parameters.

Analysis of glucose kinetics (secondary outcomes) was undertaken using normalized glucose values, to remove variation that may be caused by inter-individual differences in baseline glucose concentrations. Glucose concentrations were normalized by subtraction from the value measured before the study meal. BHB and glucose incremental

Area Under the Curve (iAUC) was calculated in GraphPad Prism 9 for Mac (version 9.1, GraphPad Software, San Diego, CA) using the trapezoid method, total iAUC is reported for BHB and net iAUC is reported for glucose as it accounts appropriately for excursions above and below the baseline (secondary outcomes).

Composite symptom scores for each subject were analyzed using unpaired t-test for between formulation and between serving size differences (exploratory outcomes). As the number of symptom reports was low, individual symptom types are not broken out by serving size.

All statistical analyses were conducted using GraphPad Prism (Version 10 for Mac OS X, Boston, USA). Data is shown as mean (SD). All p-values are two-sided and considered significant at the < 0.05 level. Analyses were performed based on observed data, any value where the ketone meter read “LO” (below limit of detection) was recorded as 0 in the data input sheet, other missing values were not imputed. All data was first tested for normality, and parametric (paired or unpaired t-test, or two-way ANOVA) or non-parametric (Mann-Whitney Test) methods with post-hoc Dunnett’s or Šídák’s multiple comparison tests were used as appropriate.

## Results

All serving sizes and formulations of BO-BD delivered nutritional ketosis (≥ 0.5 mM, indicated by dotted line) based on the mean group values from 30 mins after ingestion until the end of the study (**Figure 2A and C**). There was a significant time, subject, serving size and interaction effect on BHB for both BO-BD formulations (all p < 0.005). BHB concentrations were significantly higher than baseline at all timepoints post ingestion for both serving sizes of both conditions, expect for 60- and 90-mins post-ingestion of 12.5 g BO-BD powder. For the beverage formulation, there was a consistently greater BHB concentration in the 25 g condition vs 12.5 g condition from 1.5 h until 3 h post-ingestion (**Figure 2A**). For the powder formulation, there was no significant difference in BHB concentration between servings at any time point (**Figure 2C**).

**Figure 2:**
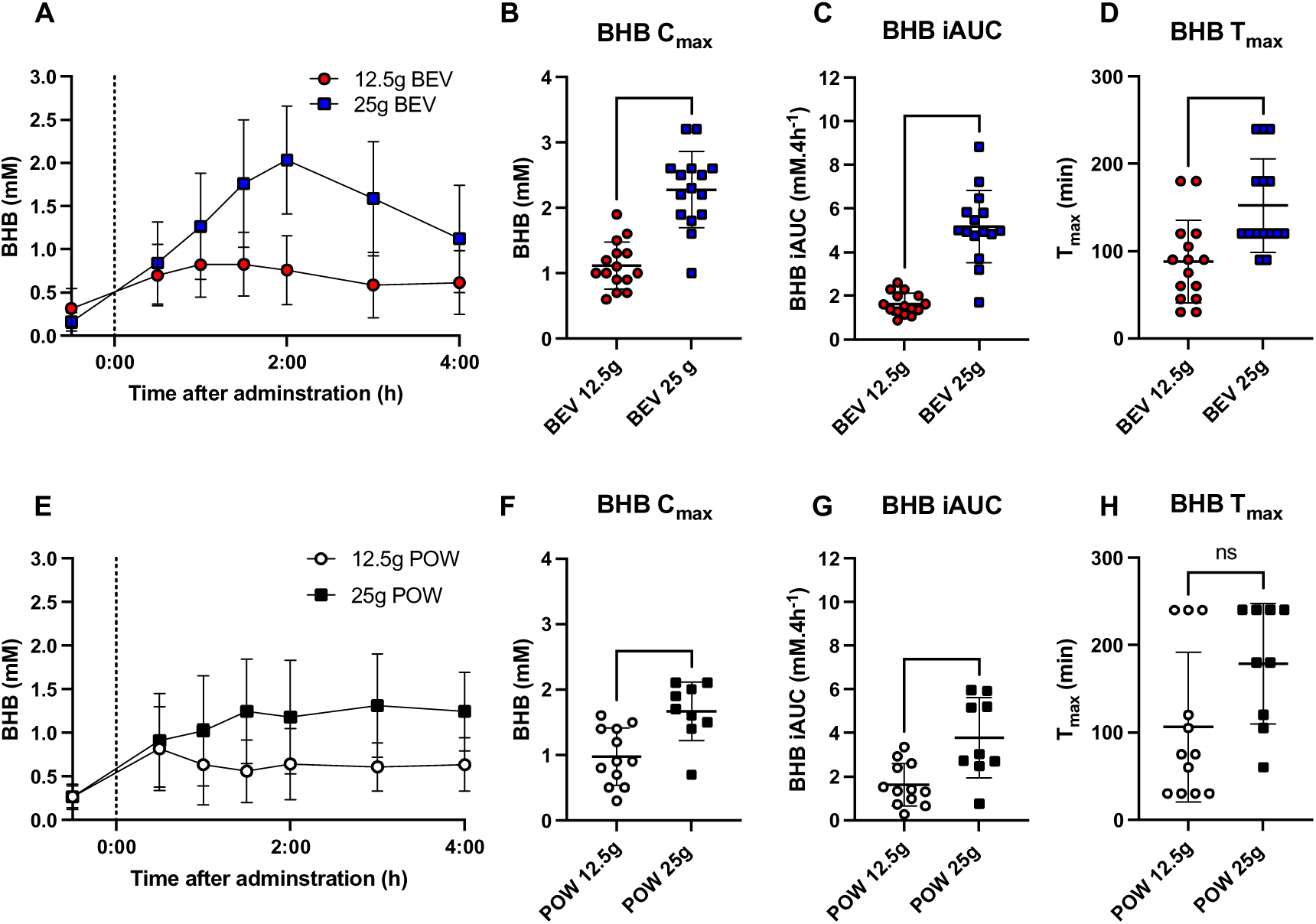
Blood BHB and glucose kinetic parameters after healthy older adults consumed 12.5 or 25 g of BO-BD in either beverage (A-D) or powder formulation (E-H). **A:** Raw blood BHB concentrations over the study after consumption of the beverage formulation; time of product consumption (time = 0) indicated by a dotted line. **B:** Individual peak BHB concentration (BEV). **C:** Incremental area under the curve for BHB per individual (BEV). **D:** Individual time of peak BHB concentration (BEV). **E:** Raw blood BHB concentrations over the study after consumption of the powder formulation; time of product consumption (time = 0) indicated by a dotted line. **F:** Individual peak BHB concentration (POW). **G:** Incremental area under the curve for BHB per individual (POW). **H:** Individual time of peak BHB concentration (POW). **Abbreviations:** BEV, beverage formulation; BHB, beta-hydroxybutyrate; C_max_, peak concentration; iAUC, incremental area under the curve, POW, powder formulation; T_max_, time of peak concentration. Results are shown as mean (SD); significance is shown as ** = p < 0.01; *** = p <0.001; **** = p <0.0001.

For the beverage formulation, increasing serving size from 12.5 g to 25 g significantly increased the BHB C_max_ by two-fold (1.1 (0.4) mM vs. 2.3 (0.6) mM, p < 0.0001, **Figure 2B**) and BHB iAUC by three-fold (1.6 (0.5) vs. 5.2 (1.7) mM.4h^−1^, p < 0.0001, **Figure 2C**), and delayed the T_max_ (88 (47) min to 152 (53) min, p = 0.0008, **Figure 2D**). There was no correlation between subject body weight and either BHB C_max_ (R^2^ = 0.0004, p =0.91) or BHB iAUC (R^2^ = 0.019, p = 0.46) (**Supplementary Figure 1 A and B**). When plotted by sex, there were no significant differences between sexes in BHB C_max_ or BHB iAUC at either serving size (**Supplementary Figure 1 C and D**), however there was a significant delay in T_max_ in female subjects vs male subjects in the 25 g serving group (180 (56) min vs. 120 (30) min, p = 0.03, **Supplementary Figure 1 E**).

For the powder formulation, increasing serving size from 12.5 g to 25 g significantly increased the BHB C_max_ by 1.7-fold (0.98 (0.4) mM vs. 1.67 (0.5) mM, p = 0.002, **Figure 2D**) and BHB iAUC by 1.8-fold (1.6 (1.0) vs. 3.8 (1.8) mM.4h^−1^, p = 0.003, **Figure 2E**), however the change in T_max_ did not reach significance (106 (86) min to 178 (69) min, p = 0.06, **Figure 2D**). There was no correlation between subject body weight and either BHB C_max_ (R^2^ = 0.07, p =0.26) or BHB iAUC (R^2^ = 0.02, p = 0.53) (**Supplementary Figure 1 F and 2G**). When plotted by sex, there were no significant differences between sexes in BHB C_max_, iAUC or T_max_ at either serving size (**Supplementary Figure 1 H, I and J**).

Analysis of data from subjects who consumed the 12.5 g serving size of the powder and beverage formulation demonstrated no significant differences in BHB C_max_, (POW = 0.94 (0.44) mM; BEV = 1.09 (035) mM, p = 0.299), iAUC (POW = 1.62 (0.98); BEV = 1.62 (0.45) mM.4h^−1^, p = 0.99) or T_max_ (POW = 106 (86) min; BEV = 90 (51) min, p = 0.85) (**Figure 3 A, B and C**). Subjects who consumed 25 g servings as powder and beverage formulation demonstrated no significant differences in BHB iAUC (POW = 3.77 (1.84); BEV = 5.09 (1.92) mM.4h^−1^, p = 0.13), or T_max_ (POW = 178 (69) min; BEV = 163 (54) min, p = 0.54), but C_max_ (POW = 1.67 (0.44) mM; BEV = 2.19 (0.64) mM, p = 0.035) was significantly greater with the beverage formulation vs the powder (**Figure 3 D, E and F**).

**Figure 3:**
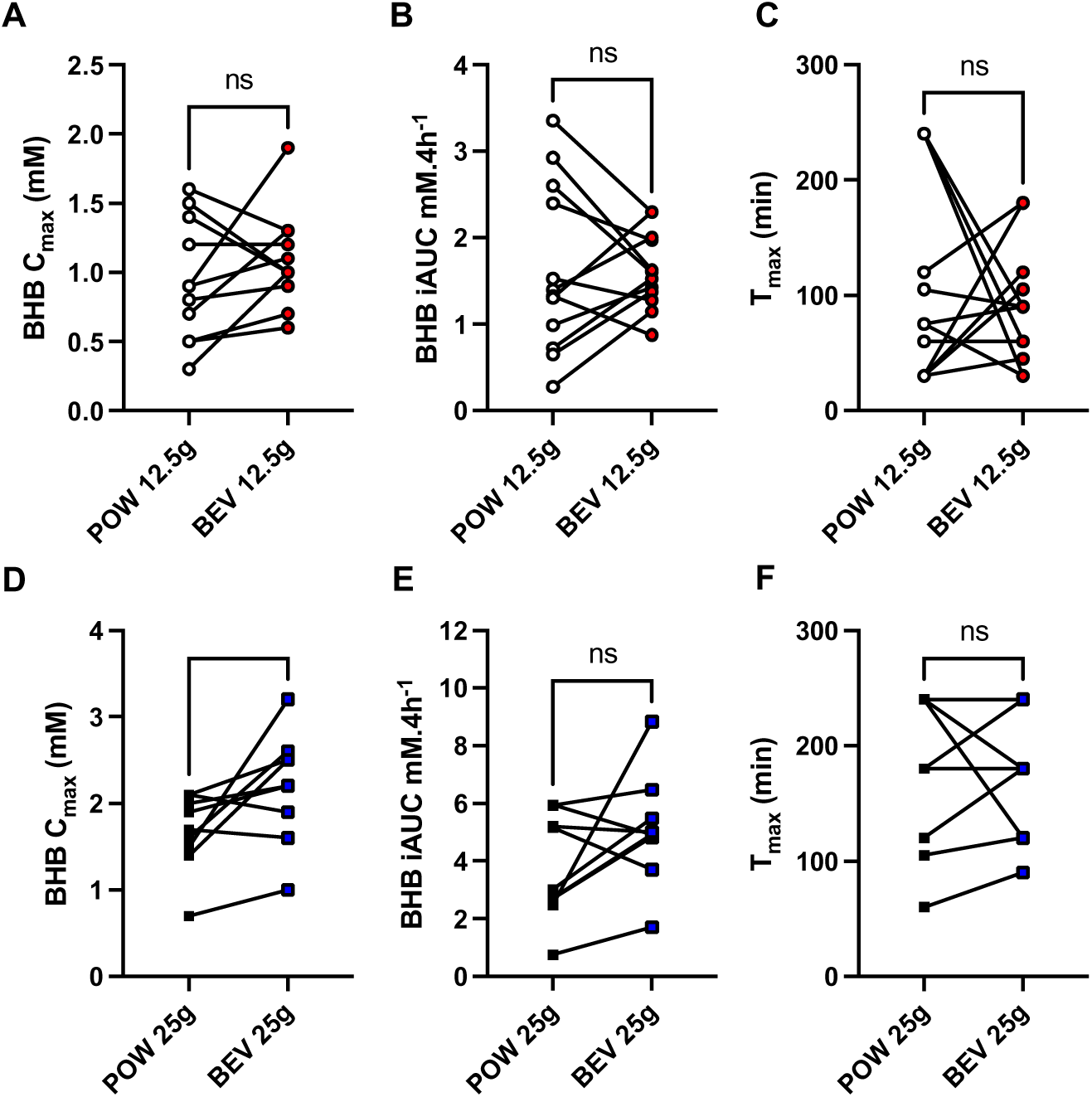
Blood ketone kinetic parameters after healthy older adults consumed 12.5 (n = 12) (**A-**C) or 25 g (n = 9) (**D-F**) of BO-BD. **A:** Peak BHB concentrations for each formulation per individual (BEV). **B:** Incremental area under the curve for BHB for each formulation per individual (BEV). **C:** Time of peak concentration for each formulation per individual (BEV). **D:** Peak BHB concentrations between formulations per individual (POW). **E:** Incremental area under the curve for BHB between formulations per individual (POW). **F:** Time of peak concentration between formulations per individual (POW). **Abbreviations:** BEV, beverage formulation; BHB, beta-hydroxybutyrate; C_max_, peak concentration; POW, powder formulation; iAUC, incremental area under the curve; T_max_, time of peak concentration. Significance is shown as * = p < 0.05.

Glucose concentration increased from baseline to 30 mins after consumption of the carbohydrate-rich standard meal with study product with both serving sizes of both BO-BD formulations (**Figure 4 A and C**). There was a significant time (p < 0.0001), serving size (p < 0.0001) and subject (p = 0.021) effect in the beverage condition, and of time (p < 0.0001), and serving size (p < 0.0001) in the powder condition. Post hoc comparisons of data from both formulations found no single time points where glucose concentration was significantly different between 12.5 g and 25 g serving groups. The net glucose iAUC was lower (i.e., smaller post-meal upward excursion, and larger post-meal reduction) in the 25 g group vs 12.5 g group (34.9 (47.1) mg/dL.4h^−1^ vs −13.4 (54.6) mg/dL.4h^−1^, p = 0.023 **Figure 4 B**) for the beverage formulation, but was not significantly different in between serving groups consuming the powder formulation (14.6 (21.8) mg/dL.4h^−1^ vs −2.9213.4 (13) mg/dL, p = 0.0 **Figure 4 D**)

**Figure 4:**
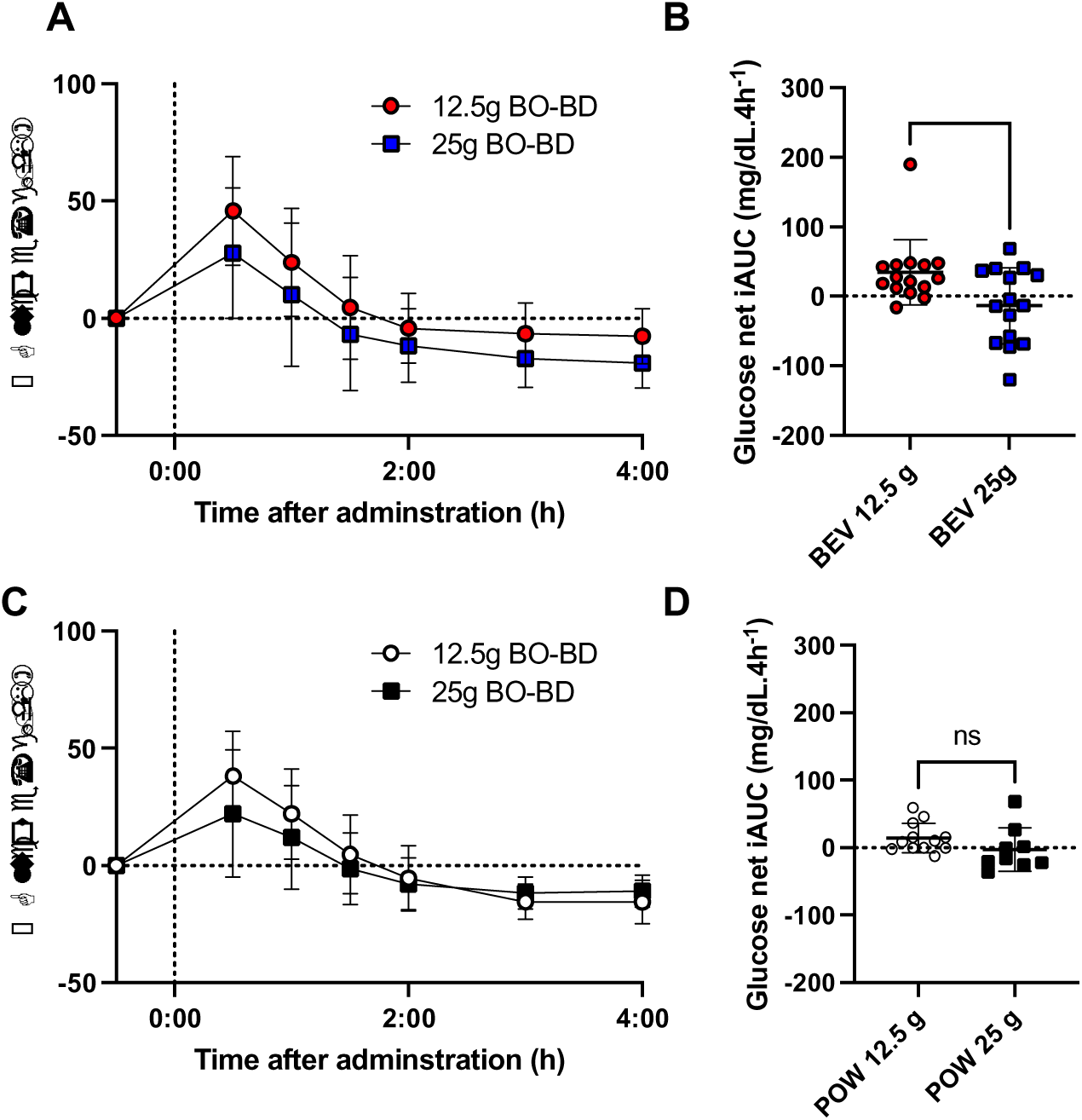
Blood glucose kinetic parameters after healthy older adults consumed 12.5 or 25 g of BO-BD formulated as beverage (n = 15 per serving group), or powder (12.5 g n = 12, 25 g n = 9). **A:** Change in glucose concentrations from baseline over the study after consumption of the beverage formulation; time of product consumption (time = 0) indicated by a dotted line. **B:** Glucose net iAUC (BEV). **C:** Change in glucose concentrations from baseline over the study after consumption of the powder formulation; time of product consumption (time = 0) indicated by a dotted line. **D:** Glucose net iAUC (POW). **Abbreviations:** BEV, beverage formulation; iAUC, incremental area under the curve; POW, powder formulation. Results are shown as mean (SD); significance is shown as * = p < 0.05.

All study products were well tolerated. The composite scores were not significantly different between either serving size given matched formulation, or between formulation giving matched serving size (median (range); BEV 12.5 g = 0 (0-2); BEV 25g = 0 (0 – 4); POW 12.5 g = 0 (0-2); POW 25g = 0 (0 – 9)). For the beverage formulation, 21 (70.0%) subjects did not report any symptoms. N = 9 subjects reported 14 mild symptoms, and 1 subject reported 1 moderate symptom of gas (**Table 3**). No subjects reported severe symptoms. For the powder formulation, n = 13 (56.5%) subjects did not report any symptoms. N = 10 subjects reported 11 mild symptoms (**Table 3**). N = 2 (8.7%) subjects reported 4 moderate or severe symptoms, including abdominal cramping, diarrhea, and dizziness (**Table 3**). All symptoms were transient and resolved before the end of the test day; there were no other adverse or serious adverse events during these test days.

**Table 3.** Tolerance of Study Beverage.

| *Symptom* | *Mild* |  | *Moderate* |  | *Severe* |  |
| --- | --- | --- | --- | --- | --- | --- |
|  | BEV | POW | BEV | POW | BEV | POW |
| Gas | 2 | 1 | 1 | 0 | 0 | 0 |
| Nausea | 0 | 2 | 0 | 0 | 0 | 0 |
| Vomiting | 0 | 0 | 0 | 0 | 0 | 0 |
| Abdominal Cramping | 0 | 0 | 0 | 2 | 0 | 0 |
| Stomach Rumbling | 3 | 0 | 0 | 0 | 0 | 0 |
| Burping | 3 | 6 | 0 | 0 | 0 | 0 |
| Reflux | 1 | 0 | 0 | 0 | 0 | 0 |
| Diarrhea | 1 | 0 | 0 | 0 | 0 | 1 |
| Headache | 0 | 0 | 0 | 0 | 0 | 0 |
| Dizziness | 4 | 2 | 0 | 1 | 0 | 0 |

**Abbreviations:** BEV, beverage formulation; POW, powder formulation.

## Discussion

This study aimed to evaluate the tolerability of, and blood ketone and glucose response to two serving sizes and two formulations of BO-BD beverage in adults 65 years or older. Both beverage and powder formulations of BO-BD at 12.5 g and 25 g serving sizes were acutely well tolerated and induced a state of nutritional ketosis (blood BHB ≥ 0.5 mM). As expected, BHB C_max_ and iAUC both increased significantly with increasing serving size of both formulations. The powder and beverage formulations had largely similar BHB kinetics, apart from a lower BHB C_max_ for powder vs beverage at 25 g. Our findings suggest that both beverage and powder formulations of BO-BD beverages are well tolerated and effective at increasing circulating ketone concentrations in older adults.

This study, examining the effects of blood ketone and glucose responses following exogenous ketone consumption exclusively in adults aged 65 years or older, addresses several notable gaps in the literature. Firstly, although there are several descriptions of the ketogenic effect of a closely related fatty acid ketone ester, bis hexanoyl (R)-1,3 butanediol (BH-BD) [40, 46, 48], this is only the second study of BO-BD. Secondly many studies have addressed exogenous ketone kinetics in younger adult populations [40, 46, 48, 49], but no study has solely recruited older adults as we have here.

BO-BD is closely structurally related to BH-BD, being different only in the length of the two fatty acid chains that make up the ester: BH-BD has six carbons per chain, and BO-BD has eight carbons per chain. BO-BD is being used here in preference to BH-BD as it has improved organoleptic properties which may ease translation into longer term studies [40]. The only available study that directly compared the two ketone esters in younger adults (mean age 48 y, range 30 – 65 y) found similar BHB, butanediol and fatty acid kinetics with both 12.5 and 25 g servings of BH-BD and BO-BD [40]. Previous studies that have administered 12.5 and 25 g of BH-BD formulated as a beverage report a BHB C_max_ of 0.8 – 1.1 mM for 12.5 g servings and ~1.7 mM for 25 g servings [40, 46, 48, 50]. Only one study has examined the kinetics of a powdered BH-BD formulation, and reported a BHB C_max_ of 0.8 and 1.7 mM for 12.5 and 25 g serving sizes respectively [48]. The results of the present study align with these previous studies of BH-BD, further confirming largely similar ketogenic efficacy between BO-BD and BH-BD.

Age could be a factor that influences blood ketone responses to ketone ester ingestion. Two studies of ketone body metabolism in rodents found that increasing age could both increase ketogenic capacity in response to a ketogenic diet [37], and increase the blood responses to exogenous ketone administration [36]. Whilst this study did not include a younger cohort, given the similar testing conditions used in previous studies of both BO-BD and BH-BD, where 12.5 and 25 g servings of ketone ester were given with a meal in younger adult cohorts, comparing data may provide clues to a possible age effect. We completed a post-hoc exploratory analysis of the data in this study (mean age 77 y) compared to our data from our most recent study of 12.5 and 25 g of BO-BD beverage taken with a standard meal (mean age 48 y). We found that for the 25 g dose group, both BHB C_max_ and BHB 4h iAUC were significantly greater in the older vs. younger cohort (BHB C_max_ old = 2.3 (0.6) mM, young = 1.8 (0.5) mM, p = 0.0092; BHB 4h iAUC old = 5.2 (1.6) mM.h^−1^, young = 3.7 (0.8) mM.h^−1^ p = 0.0009, **Supplementary Figure**). This analysis supports that there may be an effect of age to increase blood ketone responses to ketone drinks in humans, similar to the findings from the rodent studies; a larger study to investigate this relationship in more detail is currently underway (**NCT05924295**). Importantly no subject reached ketone concentrations indicative of pathological ketoacidosis (> 10 mM).

Despite all subjects reaching the accepted threshold for ketosis of 0.5 mM, we did observe a high variability in individual blood ketone responses between subjects. This study was not designed to elucidate the drivers of intra-individual variability, but we believe that factors that could have an impact may include differences in standardized meal consumption, caffeine consumption, and individual differences in digestion and metabolism. To improve the tolerance of the study products, participants were only instructed to eat until comfortably full, and differences in the amount eaten could feasibly have led to BHB or glucose response variability. It should be noted that we previously found that meal consumption does not blunt the ketogenic effect of BH-BD [46]. We also allowed subjects who were habitual caffeine users to consume caffeine, which has been shown to slightly alter blood ketone responses to ketogenic drinks [51]. Given the high variability, it may be useful to monitor ketosis on an individual basis. Blood ketone concentration could be a key determinant of functional outcomes, such as an increase in cardiac and cognitive function which are seen with higher ketone concentrations [52–54], and variability may necessitate dosing adjustments in order to exceed a threshold BHB concentration.

Exogenous ketone consumption consistently decreases fasted and post prandial blood glucose concentrations [55]. In line with this, our results show a serving size dependent decrease in glucose net iAUC with BO-BD consumption after a carbohydrate rich meal. However, without a placebo group, and without a full crossover design to account for individual variations in metabolism we cannot definitively conclude that BO-BD consumption lowered postprandial blood glucose based on our data. Previous work with BH-BD does not include non-ketone ester controls, so the effect of fatty acid based ketone esters on glucose is poorly defined.

This study has some limitations. First, it is a crossover study of formulations but not of doses, however, there were no significant differences in demographics between the 12.5 g and 25.0 g group. Second, there was no placebo control, but this would not have been feasible within the size constraints of the larger study and the primary analysis was change of blood measures from pre-consumption baseline. Third, this study was undertaken in relatively healthy adults 65 years of age and older. It remains to be determined if these findings apply to older adults with more complex or serious health conditions. Fourth, this study was not blinded, however investigator and participant knowledge of product consumption would not be expected to impact the metabolic measures (blood BHB and glucose) used and participants would have had no expectation of which formulation was more likely to cause tolerability symptoms. Finally, in both powder and beverage 25 g serving groups, blood BHB concentrations were not all returned to baseline by the end of the test day, which means the iAUC has not captured the full ketogenic effect of the study products. The length of test day was decided based on past experience with similar study products and practicality, we plan to increase sampling duration in future work.

In conclusion, both beverage and powdered formulations of the novel ketone ester BO-BD is well tolerated and induce nutritional ketosis independently of a ketogenic diet or fasting in healthy adults 65 years of age or older. The results of this study provide evidence to inform serving size selection for translation of exogenous ketone ester supplementation to target aging biology in older adults, as well as a basis for further research regarding ketone metabolism across a variety of ages and health statuses.

## Author Contributions

Conceptualization, B.J.S, J.C.N; methodology, B.J.S, J.C.N; formal analysis, B.J.S; investigation, B.J.S, E.B.S, C.S, S.R.D, S.P, W.S-M, L.A, M.Y, J.M,; data curation, B.J.S, E.B.S, C.S, S.R.D, S.P; writing—original draft preparation, B.J.S, E.B.S.; writing—review and editing, C.S, S.R.D, S.P, J.C.N; visualization, B.J.S, E.B.S; project administration, B.J.S, J.C.N.; funding acquisition, B.J.S, J.C.N. All authors have read and agreed to the published version of the manuscript.

## Author Declarations

B.J.S. has stock in H.V.M.N Inc, and stock options in Selah Therapeutics Ltd, BHB Therapeutics (Ireland) Ltd., and Juvenescence Ltd. J.C.N. has stock options in Selah Therapeutics Ltd and BHB Therapeutics (Ireland) Ltd. J.C.N and B.J.S. are inventors on patents related to the use of ketone bodies. B.J.S and J.C.N have disclosed those interests fully to The Buck Institute and Taylor & Francis, and have in place an approved plan for managing any potential conflicts arising from this arrangement. All other authors have no competing interests.

## Data Availability Statement

The data presented here may be available upon reasonable request from the corresponding author and in accordance with intellectual property considerations.

## Institutional Review Board Statement

This study was conducted according to the guidelines of the Declaration of Helsinki (2004) and registered on a registry NCT05585762. The parent study was approved by Advarra IRB on September 28^th^ 2022, the amendment that added the powder kinetics visit was approved on October 18^th^ 2023

---

**Supplementary Figure 1:**
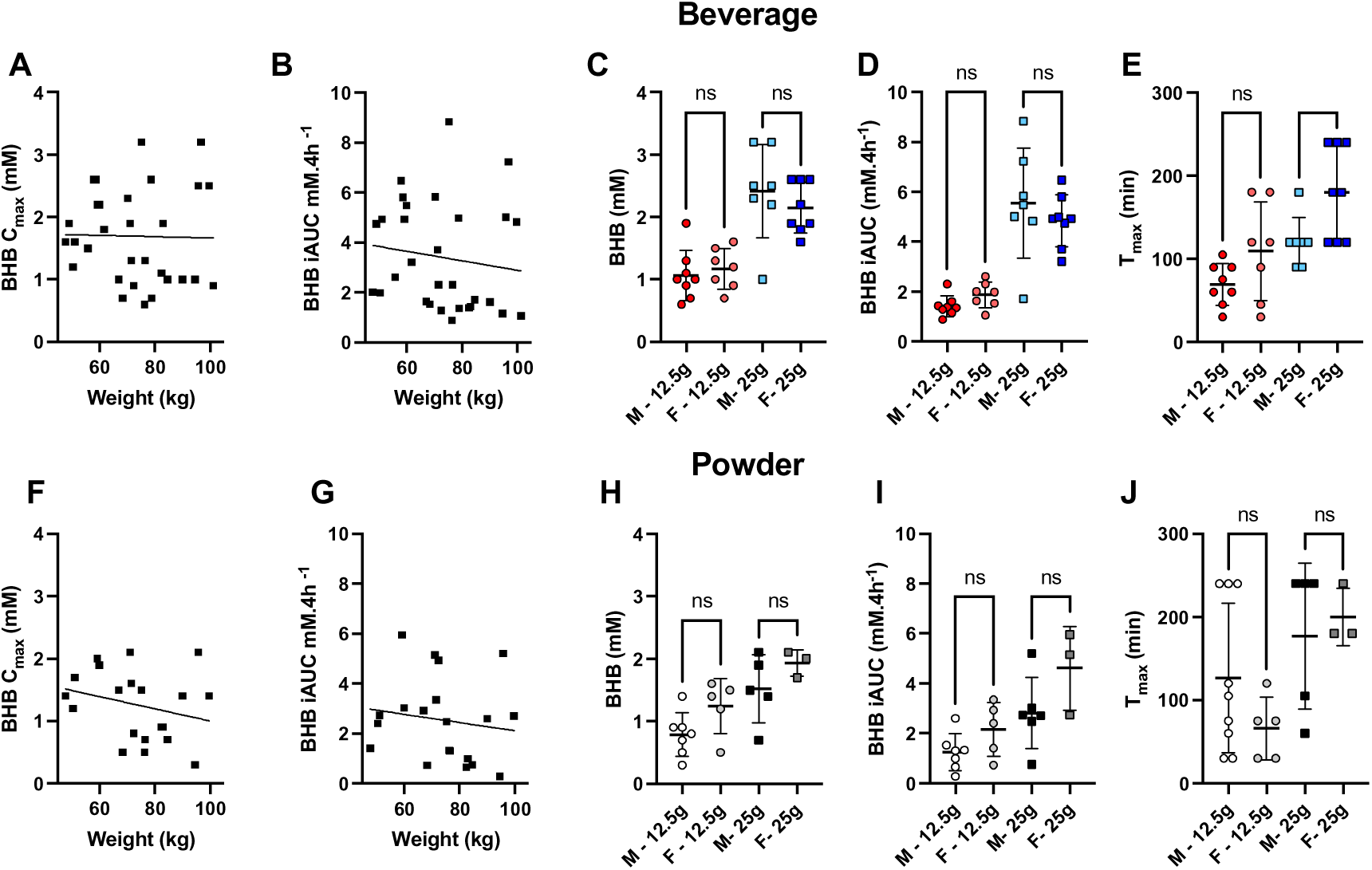
Blood ketone kinetic parameters after healthy older adults consumed 12.5 g (n = 12) or 25 g (n = 9) of BO-BD. **A:** Peak BHB concentrations compared to weight for the beverage formulation. **B.** Incremental area under the curve for BHB compared to weight for the beverage formulation. **C.** Peak BHB concentrations between males and females, by treatment group, for the beverage formulation. **D.** Incremental area under the curve for BHB between males and females, by treatment group, for the beverage formulation. **E.** Time of peak concentration of BHB between males and females, by treatment group, for the beverage formulation. **F:** Peak BHB concentrations compared to weight for the powder formulation. **G.** Incremental area under the curve for BHB compared to weight for the powder formulation. **H.** Peak BHB concentrations between males and females, by treatment group, for the powder formulation. **I.** Incremental area under the curve for BHB between males and females, by treatment group, for the powder formulation. **J.** Time of peak concentration of BHB between males and females, by treatment group, for the powder formulation. **Abbreviations:** BHB, beta-hydroxybutyrate; C_max_, peak concentration; iAUC, incremental area under the curve; T_max_, time of peak concentration; F, female; M, male. Significance is shown as * = p < 0.05.

**Supplementary Figure 2:**
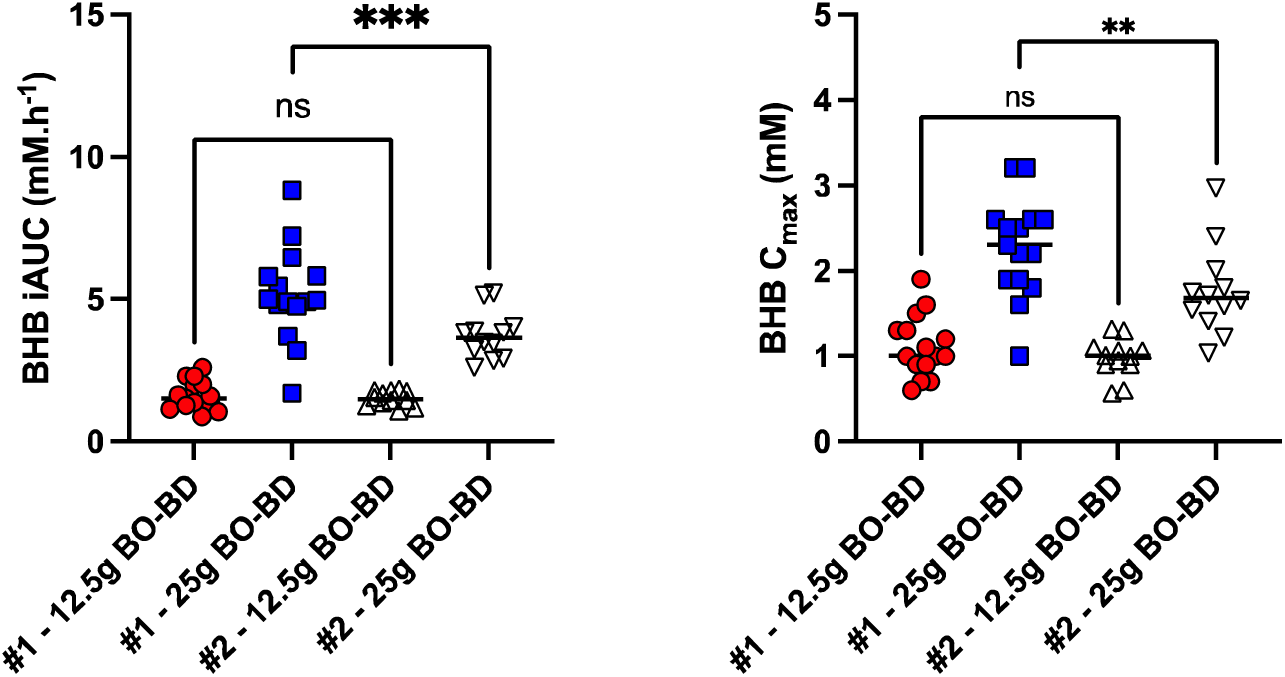
Blood ketone kinetic data from two studies where healthy older adults, mean age 76 y (#1) and 46 y (#2), consumed 12.5 g or 25 g of BO-BD beverage. **A.** Incremental area under the curve for BHB for both studies. **B:** Peak BHB concentrations for both studies. **Abbreviations:** BHB, beta-hydroxybutyrate; BO-BD, bis-octanoyl(R)-1,3-butanediol; C_max_, peak concentration; iAUC, incremental area under the curve; #1, the present study; #2, Stubbs, et al. 2023 [40]. Significance is shown as ** = p < 0.001, *** p < 0.0001.

